# Prevalence of Migraine and its associated factors among medical students of Bangladesh: A cross sectional study

**DOI:** 10.1101/2021.09.04.21263129

**Authors:** Abdur Rafi, Tasnim Shahriar, Yeasin Arafat, Abhishek Karmaker, Showsan Kabir Chowdhury, Benazir Jahangir, Meherab Hossain, Mahamoda Sultana, Md. Golam Hossain

## Abstract

**Introduction:** Medical students are vulnerable group to migraine, one of the most common type of headache worldwide. The aim of the present study was to determine the prevalence of migraine and related disability among medical students of Bangladesh.

**Methods:** This cross-sectional study was conducted among 1327 students from six medical colleges Bangladesh during March 2021 through a self-administered online survey. ID Migraine™ scale and MIDAS scale were used to screen migraine and migraine related disability respectively. Frequency distribution, and Chi-square test, t-test along with multiple logistic regressions model were used to determine the prevalence and associated factors of migraine respectively.

**Results:** The overall prevalence of migraine among the participants was 19%. The prevalence was higher among females (27%) than males (8%). Female sex (aOR 4.11, 95% CI 2.79-6.03) and poor sleep quality (aOR 2.07, 95% CI 1.48-2.91) were identified as independent risk factors of migraine. More than 90% migrainures reported to suffer from moderate to severe headache. Nausea was most commonly reported associated symptom (83.5%) followed by photophobia (72%) and vomiting (53%). Self-reported mental stress (55%), irregular sleep (49%), noise (30.5%), and usage of electronic device (30.5%) were most commonly reported triggering factor of migraine attack. More than half of the sufferers reported severe migraine related disability (MIDAS score ≥ 21).

**Conclusions:** The prevalence of migraine among medical students of Bangladesh is alarmingly high. Frequent migraine attacks and severe intensity of headache cause a substantial level of disability among the sufferers. Cautious avoidance of the triggering factors through appropriate interventions and prophylactic medication can mitigate the negative impact of migraine as well as improve the quality of life.

## Introduction

Migraine is a disabling neurological disorder characterized by localized, intense, throbbing or pulsing sensations in the head, often accompanied by nausea, vomiting, photophobia and/or phonophobia (1). It is one of the most common type of headache affecting more than 15% people worldwide (2) and considered as the eighth most burdensome disease globally (3). It is more prevalent among younger age group (4,5) resulting in hampered the quality of life and productivity a lot (6). For example, it was estimated that migraine caused a loss of 112 million days of work or school a year in US and 25 million days in UK (7). Despite these huge burden, migraine often remains underdiagnosed and untreated which makes the situation more grave (8).

Few evidences from South Asian countries showed variety in migraine prevalence ranging from 14 to 30% in India (9,10), 38% in Pakistan (11), and 35% in Nepal (12). However, there is lack of evidence on the epidemiology and characteristics of migraine as well as its effect on the day to day life of the patients in Bangladesh. A study reported that almost 26% of all headaches in Bangladesh is due to migraine (13).

Migraine attacks are triggered by a wide range of factors. Psychological stress, smoking, pattern of sleep, weather change, missing a meal, bright light, certain food and alcohol consumption etc. are considered as major triggering factors (14–16). It is evidenced that young adults are more prone to these factors which makes them vulnerable to frequent migraine attacks. Especially, medical students are identified as a more vulnerable group for developing migraine worldwide compared to other non-medical students because of the physical and psychological stressors present in their academic life (17–19). Prevalence of migraine is reported as higher among medical students in different countries ranging from 26 to 52% (12,20,21) compared to the general population (14 to 16%) (3) or other faculty students (10 to 18%) (22). This high prevalence of migraine impairs the academic performance of the students and also decrease the quality of life of the sufferers (23,24). So, it is important to explore the epidemiology and characteristics of migraine headache among the medical students as well as to mitigate the triggers to uphold the quality of life of the patients.

Despite being a significant issue, there is very little evidence reporting the prevalence, characteristics and factors associated with migraine among the medical students of Bangladesh. A study including a small sample reported that the prevalence of migraine was around 17% among the medical students of Bangladesh (25). However the study did not revealed the characteristics of migraine and factor associated with it. Hence, the present study aimed to find out the prevalence and characteristics of migraine and its associated factors among the medical students of Bangladesh.

## Methods

### Study Design and Participants

The study was a cross-sectional study conducted among the students of three government medical colleges (Khulna Medical College, Khulna, Sir Salimullah Medical College, Dhaka, and Magura Medical College, Magura) and three private medical colleges (Z. H. Sikder medical college, Dhaka, Central International Medical College, Dhaka, and Enam medical College, Dhaka) of Bangladesh. Sample size was calculated from the following formula: 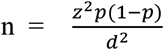, where, z = 1.96 considering the 95% confidence level, p = assumed prevalence of migraine among medical students, and d = precision of error for the prevalence estimate. A previous study reported the prevalence of migraine among medical students as 17.4 % (25). Using this information the calculated sample size was 1823. Assuming 10% non-response rate we approached 2000 medical students. Convenience sampling using an online survey was used to include participants.

### Data collection procedure

Data collection was done using a self-administered online survey form created in Google Forms. The questionnaire was reviewed by two independent consultant neurologists for content validity. Then it was pre-tested among 30 medical students who were not included in the final analysis. The survey was created and executed in English, as the medium of study was English in the medical colleges of Bangladesh. The survey link was circulated among the students after their online lecture with a request to fill up immediately. Login with email was mandatory to avoid the repetitive input of data. But we did not collect any email address to maintain the anonymity of the study. The study was conducted following the Checklist for Reporting Results of Internet ESurveys (CHERRIES) guidelines(26). Finally, a total of 1327 medical students completed the full survey.

### Data collection instrument

The survey questionnaire comprised of three parts: (i) socio-demographics, lifestyle and behavioral factors related data, (ii) headache-related data, and (iii) migraine related disability, if migraine was present using the Migraine Disability Assessment Score (MIDAS).

#### Part 1: Socio-demographics, lifestyle and behavioral factors

Socio-demographic information was collected during the survey by asking questions concerning age, gender, study year, monthly family income, marital status, height, and weight. Lifestyle and behavioral factors included fast food intake (frequency per week), amount of physical exercising (days per week for at least 30 minutes a day), smoking habits (yes/no), alcohol intake (yes/no), substance abuse (yes/no), and sleep quality measured by the Pittsburgh sleep quality index (PSQI) which is an appropriate screening tool for measuring sleep dysfunction in both clinical and non-clinical samples. The PSQI score was categorized as poor (PSQI score >5) and good (PSQI score ≤5) (27).

#### Part 2: Headache related data

Participants were initially evaluated by the question “*Did you have two or more headaches in the last 3 months?*” Those who responded ‘yes” were considered as the subjects with potentially troublesome headaches and further screened using the ID Migraine™ test. The ID-Migraine™ test is a three-item self-administered screening tool, developed by Lipton et al. (2003) (28). It consists of three questions regarding problems related to migraines over the last three months: 1. *Did you feel nauseated or sick in your stomach with your headaches?* 2. *Did light bother you when you had a headache (a lot more than when you do not have headaches)?* and 3. *Did your headache limit your ability to work, study or do what you needed to do for at least 1 day?* A test-diagnosis of migraine headache is made by at least two positive responses. ID Migraine™ has been validated using the International Classification of Headache Disorders (ICHD) criteria in different studies with a pooled sensitivity of 0.84 and a specificity of 0.76 (29).

Headache related data were collected from the participants who were screened as positive for migraine. These included intensity of headache (measured on a four-point scale where 0 = no headache; 1 = mild headache; 2 = moderate headache; 3 = severe headache recommended for use in migraine research by the International Headache Society) (30), frequency of headache during the past month, associated symptoms of headache, characteristic of headache (unilateral, bilateral, pulsating, and throbbing), frequency of analgesic use during the past month, frequency of healthcare facility visit during past 12 months, migraine triggers, and family history of migraine.

#### Part 3: Migraine related disability

Disability due to migraine among the participants was assessed using the Migraine Disability Assessment Score (MIDAS). The MIDAS was primarily developed and validated in general population with migraine by Stewart et. al. (31). It is a simple and easily interpretable well validated disability screening tool and widely used among migrainure medical students (24,32,33). Disability is classified into four grades according to the MIDAS scale: little or no disability (scores range 0–5), mild disability (scores range 6– 10), moderate disability (scores range 11–20), and severe disability (scores 21 or greater).

### Statistical analysis

All statistical analyses were conducted using the SPSS version 24.0. Descriptive statistics like frequency with percentage were used for categorical variables and mean with SD (Standard deviation) were used for continuous variables. Chi-square test was used to test the difference between groups in case of categorical variables and independent t-test was used in case of continuous variables. Multiple logistic regression models including the significant variables of the bivariate analysis were used to find out the factors associated with migraine. A p-value of < 0.05 with 95% confidence interval (CI) was considered as statistically significant.

## Results

### Socio-demographic characteristics

The participants of the study aged between 17 to 25 years (mean 22, SD 2 years). More than 56% of them were female and belonged to middle (43%) or high (44%) income family. Almost 27% of the participants were overweight. The daily screen time was more than six hours of 39% of the participants. Almost 9% were smoker and 6.5% were alcohol or other substance abuser (Table 1).

**Table 1:** Sociodemographic characteristics of the participants according to migraine status (n=1327)

| Characteristics | Total | Migraine | No migraine | p-value |
| --- | --- | --- | --- | --- |
| Age | 22.1 (2.0) | 21.5 (2.1) | 22.2 (2.7) | 0.181 |
| Age (category) |  |  |  |  |
| 17-19 | 380 (28.6) | 91 (23.9) | 289 (76.1) | 0.008 |
| 20-22 | 704 (53.1) | 115 (16.3) | 589 (83.7) |  |
| 23-25 | 243 (18.3) | 43 (17.7) | 200 (82.3) |  |
| Sex |  |  |  |  |
| Female | 751 (56.6) | 203 (27.0) | 548 (73.0) | <0.001 |
| Male | 576 (43.4) | 46 (8.0) | 530 (92.0) |  |
| Study year |  |  |  |  |
| 1<sup>st</sup> | 299 (22.5) | 66 (22.1) | 233 (77.9) | 0.019 |
| 2<sup>nd</sup> | 225 (17.0) | 52 (23.1) | 173 (76.9) |  |
| 3<sup>rd</sup> | 202 (15.2) | 41 (20.3) | 161 (79.7) |  |
| 4<sup>th</sup> | 344 (25.9) | 47 (13.7) | 297 (86.3) |  |
| 5<sup>th</sup> | 257 (19.4) | 43 (16.7) | 214 (83.3) |  |
| Family income |  |  |  |  |
| Low | 180 (13.6) | 31 (17.2) | 149 (82.8) | 0.353 |
| Middle | 567 (42.7) | 99 (17.5) | 468 (82.5) |  |
| High | 580 (43.7) | 119 (20.5) | 461 (79.5) |  |
| Marital status |  |  |  |  |
| Unmarried | 1274 (96.0) | 237 (18.6) | 1037 (81.4) | 0.461 |
| Married | 53 (4.0) | 12 (22.6) | 41 (77.4) |  |
| BMI (kg/m<sup>2</sup>) |  |  |  |  |
| Overweight ($\geq 25$) | 353 (26.6) | 70 (19.8) | 283 (80.2) | 0.346 |
| Normal (18.5–24.9) | 841 (63.4) | 149 (17.7) | 692 (82.3) |  |
| Underweight ($< 18.5$) | 133 (10.0) | 30 (22.6) | 103 (77.4) | |
| Exercise frequency (days per week) |  |  |  |  |
| $\geq 3$ | 240 (18.1) | 30 (12.5) | 210 (87.5) | 0.018 |
| 1-2 | 227 (17.1) | 42 (18.5) | 185 (81.5) |  |
| $< 1$ | 860 (64.8) | 177 (20.6) | 683 (79.4) | |
| Daily screen time (hours) |  |  |  |  |
| $> 12$ | 103 (7.8) | 21 (20.4) | 82 (79.6) | 0.897 |
| 6-12 | 410 (30.9) | 75 (18.3) | 335 (81.7) |  |
| 2-6 | 723 (54.5) | 138 (19.1) | 585 (80.9) |  |
| $< 2$ | 91 (6.9) | 15 (16.5) | 76 (83.5) | |
| Smoking |  |  |  |  |
| Yes | 122 (9.2) | 11 (9.0) | 111 (91.0) | 0.004 |
| No | 1205 (90.8) | 238 (19.8) | 967 (80.2) |  |
| Alcohol |  |  |  |  |
| Yes | 68 (5.1) | 5 (7.4) | 63 (92.6) | 0.013 |
| No | 1259 (94.9) | 244 (19.4) | 1015 (80.6) |  |
| Substance abuse |  |  |  |  |
| Yes | 19 (1.4) | 0 (0.0) | 19 (100.0) | 0.134 |
| No | 1308 (98.6) | 249 (19.0) | 1059 (81.0) |  |
| Sleep quality |  |  |  |  |
| Poor (PSQI score $> 5$) | 842 (63.5) | 194 (23.0) | 648 (77.0) | $< 0.001$ |
| Good (PSQI score $\leq 5$) | 485 (36.5) | 55 (11.3) | 430 (88.7) | |

### Prevalence of migraine

A total of 249 (19%) participants were screened as positive in ID Migraine™ scale. Prevalence of migraine was higher among younger (24% in 17-19 years age group) and female participants (27% in female vs 8% in male) (Table 1).

Chi-square test showed that lack of regular exercise (less than once a week), smoking tobacco, alcohol drinking and poor sleep quality were associated with migraine (Table 1). However, in logistic regression models, it was found that female participants had four times higher risk of having migraine (aOR 4.11, 95% CI 2.79-6.03, p-value <0.001) while those who had poor sleep quality had two times higher risk than their counterpart’s (aOR 2.07, 95% CI 1.48-2.91, p-value <0.001) (table 2).

**Table 2:** Factors associated with migraine (multiple logistic regression models)

| Characteristics | aOR (95% CI) | p-value |
| --- | --- | --- |
| Age (years) |  |  |
| 17-19 | 0.84 (0.41-1.69) | 0.628 |
| 20-22 | 0.62 (0.36-1.05) | 0.080 |
| 23-25 | Ref. |  |
| Sex |  |  |
| Female | 4.11 (2.79-6.03) | <0.001 |
| Male | Ref. |  |
| Study year |  |  |
| 1<sup>st</sup> | 1.38 (0.68-2.81) | 0.369 |
| 2<sup>nd</sup> | 1.86 (0.97-3.54) | 0.060 |
| 3<sup>rd</sup> | 1.58 (0.85-2.95) | 0.148 |
| 4<sup>th</sup> | 0.92 (0.53-1.60) | 0.786 |
| 5<sup>th</sup> | Ref. |  |
| Exercise frequency (days per week) |  |  |
| ≥3 | 0.76 (0.49-1.18) | 0.227 |
| 1-2 | 1.31 (0.87-1.96) | 0.188 |
| <1 | Ref. |  |
| Smoking |  |  |
| Yes | 1.31 (0.61-2.76) | 0.481 |
| No | Ref. |  |
| Alcohol intake |  |  |
| Yes | 0.46 (0.17-1.27) | 0.138 |
| No | Ref. |  |
| Sleep quality |  |  |
| Poor (PSQI score >5) | 2.07 (1.48-2.91) | <0.001 |
| Good (PSQI score ≤5) | Ref. |  |

**Table 3:**
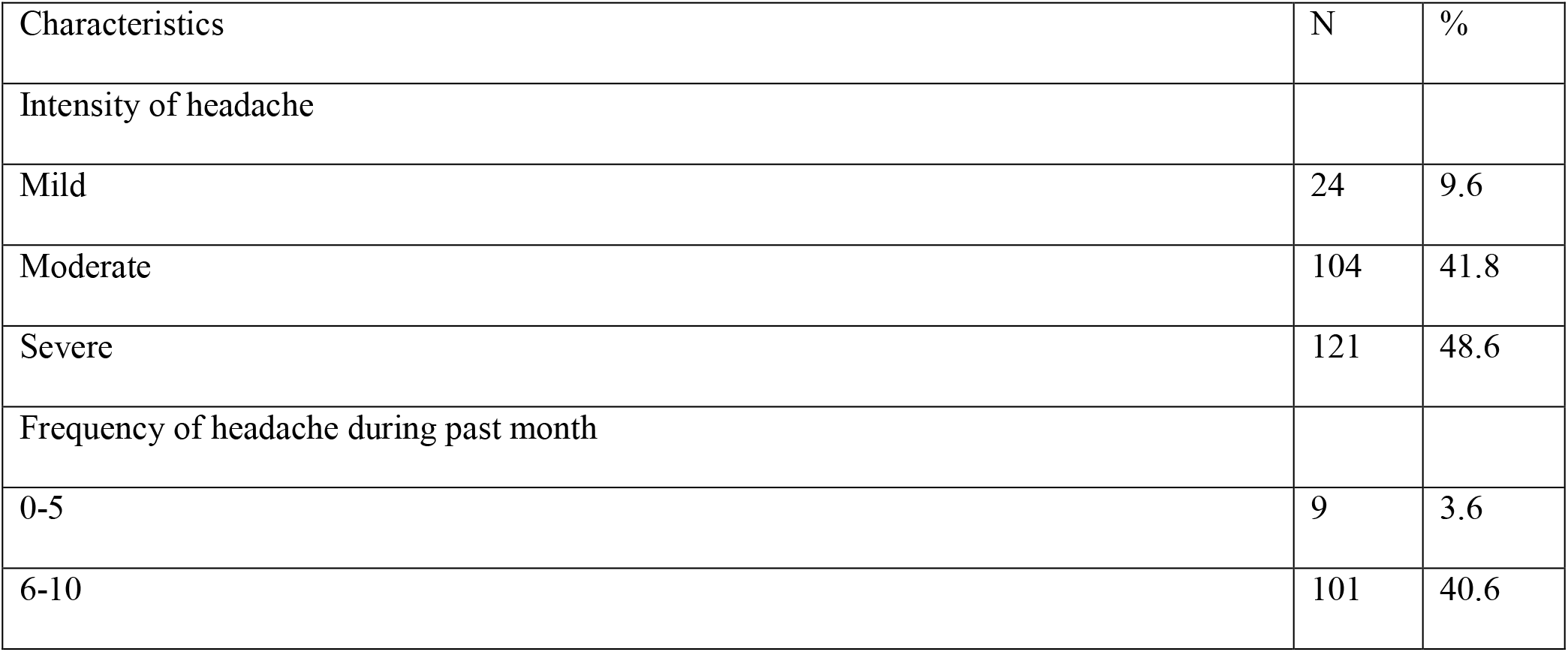
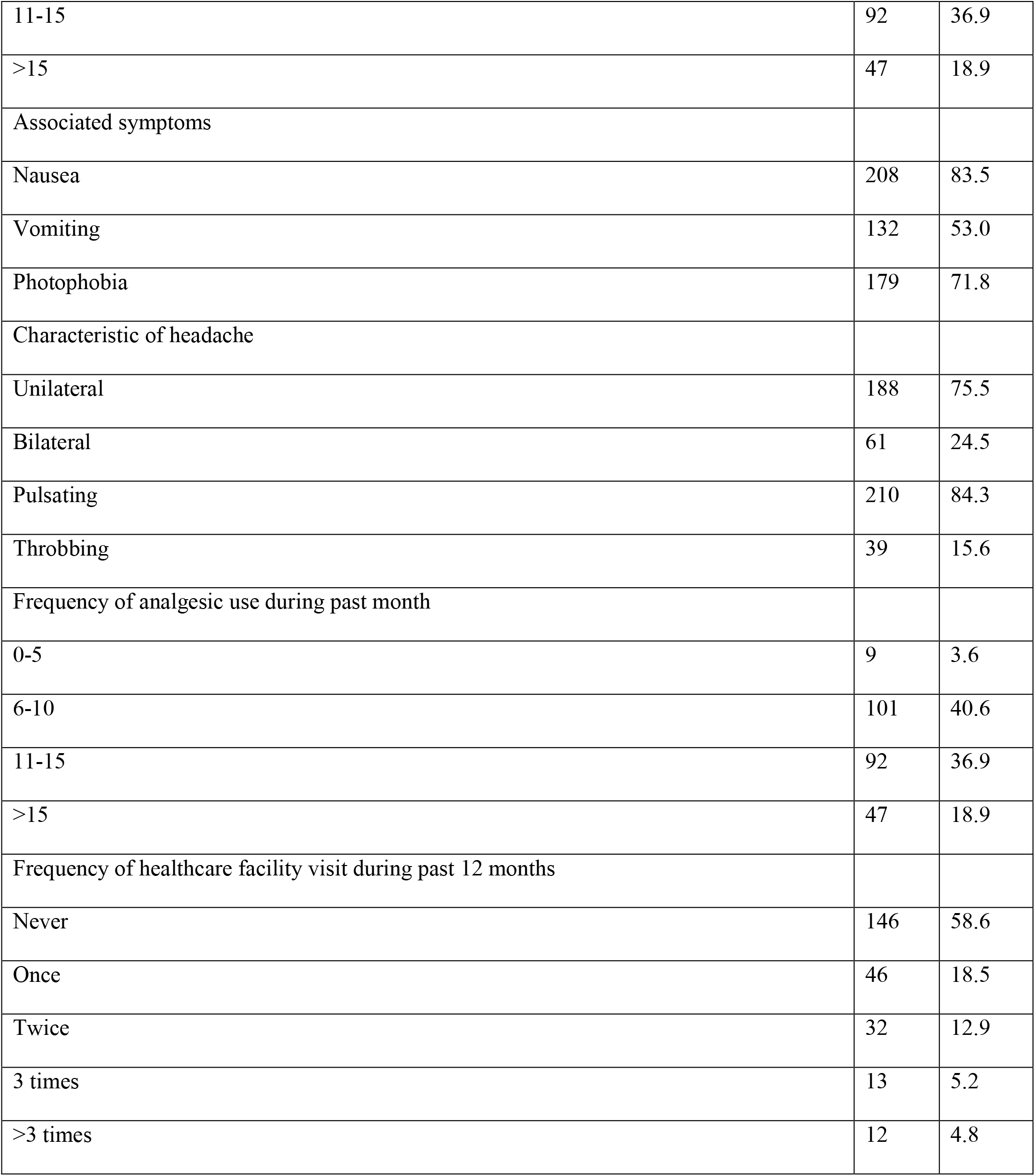
Characteristics of migraine of ID migraine headache positive participants (n=249)

| Characteristics | N | % |
| --- | --- | --- |
| Intensity of headache |  |  |
| Mild | 24 | 9.6 |
| Moderate | 104 | 41.8 |
| Severe | 121 | 48.6 |
| Frequency of headache during past month |  |  |
| 0-5 | 9 | 3.6 |
| 6-10 | 101 | 40.6 |
| 11-15 | 92 | 36.9 |
| >15 | 47 | 18.9 |
| Associated symptoms |  |  |
| Nausea | 208 | 83.5 |
| Vomiting | 132 | 53.0 |
| Photophobia | 179 | 71.8 |
| Characteristic of headache |  |  |
| Unilateral | 188 | 75.5 |
| Bilateral | 61 | 24.5 |
| Pulsating | 210 | 84.3 |
| Throbbing | 39 | 15.6 |
| Frequency of analgesic use during past month |  |  |
| 0-5 | 9 | 3.6 |
| 6-10 | 101 | 40.6 |
| 11-15 | 92 | 36.9 |
| >15 | 47 | 18.9 |
| Frequency of healthcare facility visit during past 12 months |  |  |
| Never | 146 | 58.6 |
| Once | 46 | 18.5 |
| Twice | 32 | 12.9 |
| 3 times | 13 | 5.2 |
| >3 times | 12 | 4.8 |

### Characteristics of migraine

Among 249 participants with migraine more than 90% reported to suffer from moderate to severe headache (42 and 48% respectively). Almost 56% of them had more than 10 attacks during the past month. Nausea was most commonly reported associated symptom (83.5%) followed by photophobia (72%) and vomiting (53%). More than 75% migraineurs had unilateral headache while more than 83% had pulsating type of headache (Table 2). Self-reported mental stress was most commonly reported triggering factor of migraine attack (55%) followed by irregular sleep (49%), noise (30.5%), usage of electronic device (30.5%) and physical activities (23%) (Figure 1).

**Figure 1:**
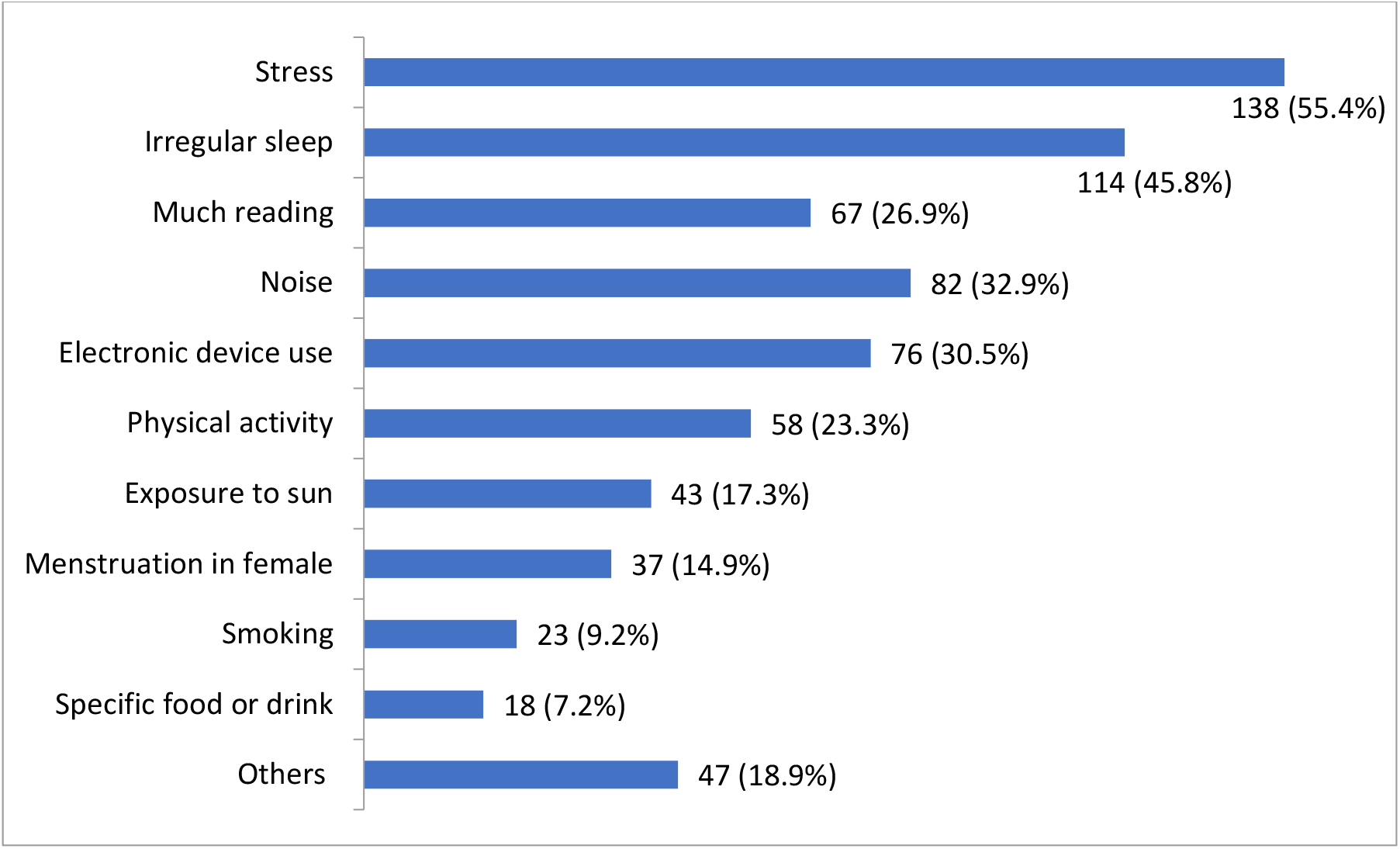
Triggers of migraine attacks reported by the ID Migraine positive participants (n=249) Migraine related disability

Only 11.6% of the participants with migraine reported little or no migraine related disability as measured by MIDAS scale (score ≤5). Almost 20% and 16% of them reported to have mild and moderate migraine related disability while more than 52% reported severe disability (MIDAS score ≥ 21) (Figure 2).

**Figure 2:**
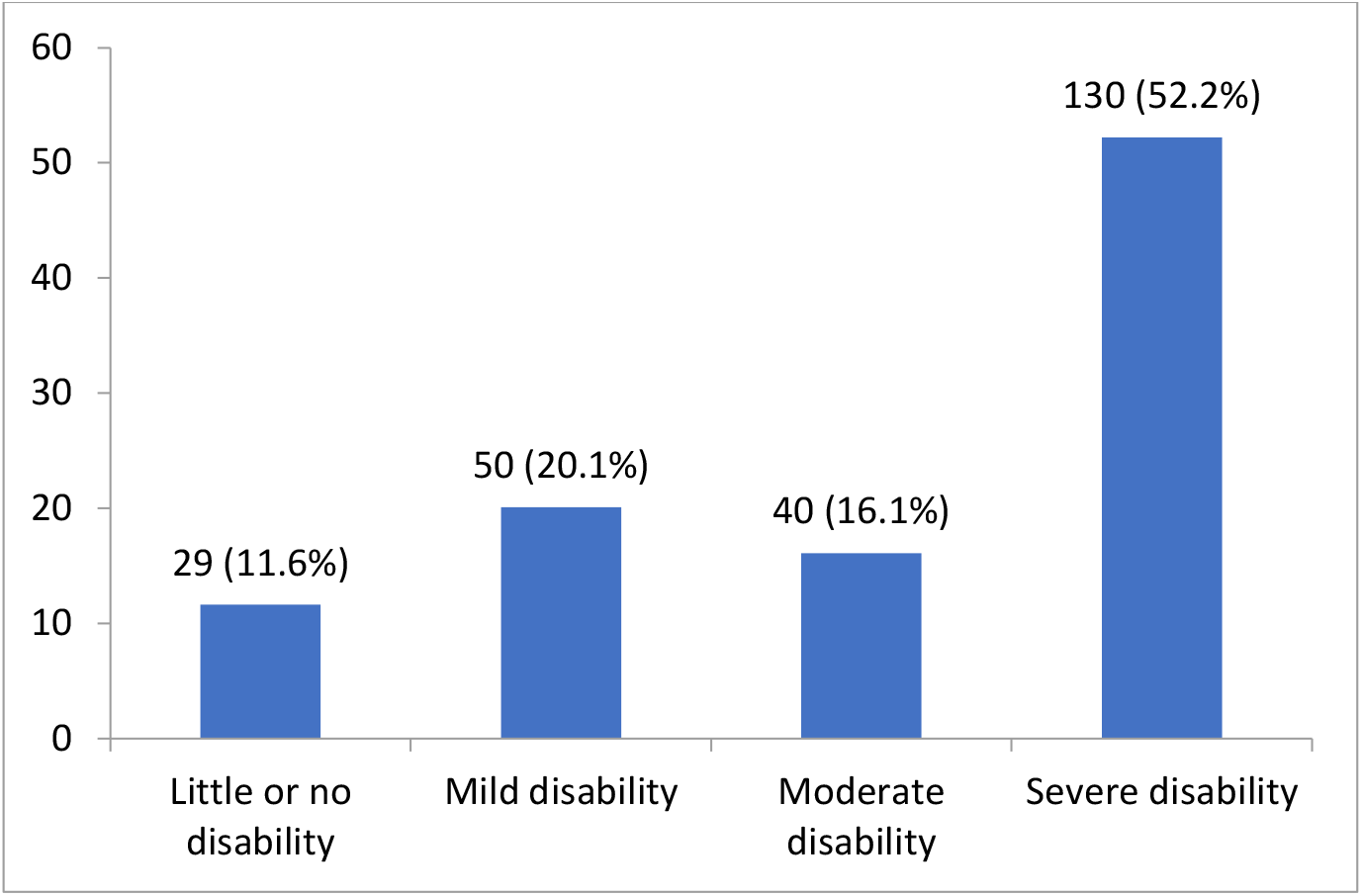
Migraine related disability reported by the ID Migraine positive participants (n=249)

## Discussion

Epidemiological studies can play important role in finding the prevalence of a disease or/and event and as well as to inform the policy makers to make the best decision for using resources in appropriate space which is important in countries with limited resources in health sector like Bangladesh. The main objective of our study was to find out the prevalence and characteristics of migraine and its associated factors among the medical students of Bangladesh. To our best knowledge, there was very few evidence on the topic in Bangladesh. In our study, the prevalence of migraine among medical students was 19% which was within the worldwide reported range among medical students. A study conducted in neighboring India reported somewhat similar statistics (prevalence of migraine among medical students 14%) (9). However some other studies reported a comparatively higher prevalence of migraine among this population group of South East Asian region such as 30% in India (10), 38% in Pakistan (11), and 35% in Nepal (12). In the studies conducted in different countries like Brazil (12%) (34), Nigeria (14%) (35), Turkey (13%) (36), and Oman (12%) reported slightly lower prevalence of migraine compared to our study. On the contrary, Saudi Arabia (24%) (37), Nairobi (34%) (38), or Kuwait (28%) (23) reported a higher prevalence of migraine. And this worldwide variation of migraine prevalence among medical students could be explained as there are different stressors and triggering factors in different universities and in different areas. Different diagnosis criteria, geographical location, socioeconomic and nutritional status, personal habits and physical activities could also be a contributing factor in variation of migraine prevalence.

Female students in the present study showed significantly higher prevalence of migraine compared to their male counterparts. Several studies also supported our finding of the higher prevalence of migraine among female participants (39–41). This phenomenon may due to hormonal influence particularly estrogen. Estrogen increases the Ca^2+^ levels and decreases Mg^2+^ level, causing an imbalance that may enhance neuronal excitation and induce migraine. It also enhances the synthesis and release of calcitonin gene-related peptide CGRP and nitric oxide, which may trigger migraine by reinforcing vasodilation with activating and transmitting pain signals to the trigeminal nerve (42).

We have found that students from first three years of their academic life had more prevalence which is also supported by a study in China (43) though a study from Saudi Arabia showed the opposite result where students from fifth year suffered most from the migraine (37). One of the plausible explanations to this difference could be the first year and newer students in our study participants face more difficulties to cope up with the newer environment and curriculum of medical study after completing their high school degree. Literature suggests that sedentary lifestyle, smoking tobacco, alcohol and substance abuse and sleep disturbance were associated with migraine (44–46). Though we found poor sleep quality as a risk factor of migraine, the others were not significantly associated in our study.

In our study mental stress was most common reported triggering factor of migraine attack. It was predictable as medical students are exposed to stress regarding their exams, high-level performance, long duration of education, and the constant responsibility to the course (28). And irregular sleep, noise, usage of electronic device, physical activities are respectively the common triggering factor next to mental stress. Some previous studies also suggested similar triggering factors for migraine attack. For example, in a study among medical students of China stress at study or work and lack of sleep were most common triggering factors for migraine (43). Similar results were also found in studies from India (10), Saudi Arabia (37) and Pakistan (11).

Most of the participants diagnosed with migraine reported to suffer from moderate to severe headache. Among them more than half of the participants had more than ten attacks during the past month. In Turkey, the average of migraine episode was 5.9 per month (36) and in Saudi Arabia it was 4.6 per month (20). This immense rate of attack and intensity of headache among migraineurs caused severe migraine related disability. Among our participants, more than half reported severe disability. This rate of disability was unusually higher than previously reported studies. In Saudi Arabia (47) and Turkey (36) severe disability was consequently 29% and 22%. This rate of disability reduces the productivity of students which can be as much as half of the overall productivity (34). To reduce this severe level of disability controlling the trigger factors along with medicine take upon counselling with physician is needed.

### Limitations

There are several limitations that are needed to be considered to interpret the findings of our study. Firstly, diagnosis of migraine wasn’t done clinically rather we used a self-administered screening tool which could affect the overall result of migraine prevalence. Moreover, recall bias of the participants is also a considerable limitation. Moreover, being an online survey we could not roll out the risk of sampling bias. However, more extensive and clinical diagnosis based studies are needed in order to understand prevalence and characteristics of migraine more precisely.

## Conclusion

Our study found that prevalence of migraine among medical students of Bangladesh was high. Female and junior class students are more frequently affected by migraine. It could potentially reduce their academic performance and impact negatively on their productivity. Triggering factors like mental stress, smoking tobacco, alcohol drinking, etc. are needed to be mitigated in order to reduce the risk of repeated migraine attacks. Early screening and management can reduce the burden of migraine.

## Ethical consideration

The present study was carried out in accordance with the ethical standard of the World Medical Association (i.e., declaration of Helsinki) as well as the guidelines of Institutional research ethics. The formal ethical approval was endorsed by the Ethical Review Committee of Rajshahi Medical College (RMC/ERC/2020-2022/198/178). Participants were well informed about the procedure and purpose of the study, and confidentiality of their information. Electronic informed consent was ensured by each of participants.

## Data Availability

Data will be available on request to the corresponding author.

## Consent for publication

Not applicable.

## Availability of data and materials

Data will be available on request to the corresponding author.

## Author Contributions

Conceptualization: Abdur Rafi, Md. Golam Hossain.

Formal analysis: Abdur Rafi, Md. Golam Hossain.

Investigation: Abdur Rafi, Tasnim Shahriar, Yeasin Arafat, Abhishek Karmaker, Showsan Kabir Chowdhury, Benazir Jahangir, Meherab Hossain, Mahamoda Sultana.

Methodology: Abdur Rafi, Tasnim Shahriar, Md. Golam Hossain.

Resources: Showsan Kabir Chowdhury, Benazir Jahangir.

Supervision: Md. Golam Hossain

Writing – original draft: Abdur Rafi, Tasnim Shahriar.

Writing – review and editing: Abdur Rafi, Tasnim Shahriar, Yeasin Arafat, Abhishek Karmaker, Showsan Kabir Chowdhury, Benazir Jahangir, Meherab Hossain, Mahamoda Sultana, Md. Golam Hossain.

## Acknowledgments

We would like to express their profound gratitude to Dr. Azizul Haque, associate professor of Rajshahi Medical College and Dr. Idris Ali, consultant of Neuromedicine department of Rajshahi Medical College Hospital for their important role in reviewing the survey instruments. We are also thankful to Farhan Jadid, Mastura Jannat Mridula and Sajib Kumar Ghosh for their voluntary contributions during the survey periods.

## References

1. Olesen J, Bes A, Kunkel R, Lance JW, Nappi G, Pfaffenrath V, et al. The International Classification of Headache Disorders, 3rd edition (beta version). Cephalalgia [Internet]. 2013 Jul 1 [cited 2020 Sep 16];33(9):629–808. Available from: https://pubmed.ncbi.nlm.nih.gov/23771276/

2. Vos T, Flaxman AD, Naghavi M, Lozano R, Michaud C, Ezzati M, et al. Years lived with disability (YLDs) for 1160 sequelae of 289 diseases and injuries 1990-2010: A systematic analysis for the Global Burden of Disease Study 2010. Lancet. 2012;380(9859):2163–96.

3. Leonardi M, Raggi A. Burden of migraine: International perspectives. Neurol Sci. 2013 May 22;34(SUPPL. 1):117–8.

4. Woldeamanuel YW, Andreou AP, Cowan RP. Prevalence of migraine headache and its weight on neurological burden in Africa: A 43-year systematic review and meta-analysis of community-based studies. Vol. 342, Journal of the Neurological Sciences. Elsevier; 2014. p. 1–15.

5. Hazard E, Munakata J, Bigal ME, Rupnow MFT, Lipton RB. The burden of migraine in the United States: Current and emerging perspectives on disease management and economic analysis. Value Heal. 2009;12(1):55–64.

6. Lipton RB, Stewart WF, Scher AI. Epidemiology and economic impact of migraine. Vol. 17 Suppl 1, Current medical research and opinion. Curr Med Res Opin; 2001.

7. TJ S, AI S, WF S, K K, J L, RB L. The prevalence and disability burden of adult migraine in England and their relationships to age, gender and ethnicity. Cephalalgia. 2003 Sep;23(7):519–27.

8. Woldeamanuel YW, Cowan RP. Migraine affects 1 in 10 people worldwide featuring recent rise: A systematic review and meta-analysis of community-based studies involving 6 million participants. J Neurol Sci. 2017;372:307–15.

9. Ray, Paul N, Hazra A, Das S, Ghosal MK, Misra AK, et al. Prevalence, burden, and risk factors of migraine: A community-based study from Eastern India. Neurol India. 2017 Nov;65(6):1280.

10. Raju S S G., Prevalence of migraine among medical students of a tertiary care teaching medical college and hospital in South India - A cross-sectional study. Natl J Physiol Pharm Pharmacol. 2018;8(9):1377.

11. khan A, khattak hammad, jamali R, rashid hina, riaz ayesha, ibrahimzai arsalan khan. PREVALENCE OF MIGRAINE, ITS COMMON TRIGGERING FACTORS AND COPING STRATEGIES IN MEDICAL STUDENTS OF PESHAWAR. KHYBER Med Univ J. 2012 Dec;4(4):187–92.

12. Manandhar K, Risal A, Steiner TJ, Holen A, Linde M. The prevalence of primary headache disorders in Nepal: a nationwide population-based study. J Headache Pain 2015 161. 2015 Nov;16(1):1–10.

13. Hussain AM, Mohit M, Ahad M, Alim M. A Study on Psychiatric Co-morbidity Among the Patients with Migraine. TAJ J Teach Assoc. 1970 Jan;21(2):108–11.

14. López-Mesonero L, Márquez S, Parra P, Gámez-Leyva G, Muñoz P, Pascual J. Smoking as a precipitating factor for migraine: A survey in medical students. J Headache Pain. 2009;10(2):101–3.

15. Nazari F, Safavi M, Mahmudi M. Migraine and its relation with lifestyle in women. Pain Pract. 2010 May;10(3):228–34.

16. Takeshima T, Ishizaki K, Fukuhara Y, Ijiri T, Kusumi M, Wakutani Y, et al. Population-Based Door-to-Door Survey of Migraine in Japan: The Daisen Study. Headache. 2004 Jan;44(1):8–19.

17. Dyrbye LN, Thomas MR, Shanafelt TD. Systematic review of depression, anxiety, and other indicators of psychological distress among U.S. and Canadian medical students. Vol. 81, Academic Medicine. Lippincott Williams and Wilkins; 2006. p. 354–73.

18. Bigal ME, Bigal JM, Betti M, Bordini CA, Speciali JG. Evaluation of the impact of migraine and episodic tension-type headache on the quality of life and performance of a university student population. Headache. 2001;41(7):710–9.

19. Smitherman TA, McDermott MJ, Buchanan EM. Negative impact of episodic migraine on a university population: Quality of life, functional impairment, and comorbid psychiatric symptoms. Headache. 2011 Apr;51(4):581–9.

20. Ibrahim NK, Alotaibi AK, Alhazmi AM, Alshehri RZ, Saimaldaher RN, Murad MA. Prevalence, predictors and triggers of migraine headache among medical students and interns in King Abdulaziz University, Jeddah, Saudi Arabia. Pakistan J Med Sci. 2017 Mar;33(2):270–5.

21. Anwar F, Bilal Sheikh A, Taher T, Iqbal Khan M, Masoom A, Khursheed A, et al. Prevalence and predictors of migraine among medical students in Karachi. J Basic Res Med Sci. 2021;8(1):19–27.

22. Wang X, Zhou HB, Sun JM, Xing YH, Zhu YL, Zhao YS. The prevalence of migraine in university students: A systematic review and meta-analysis [Internet]. Vol. 23, European Journal of Neurology. Blackwell Publishing Ltd; 2016 [cited 2020 Sep 16]. p. 464–75. Available from: https://onlinelibrary.wiley.com/doi/abs/10.1111/ene.12784

23. Al-Hashel JY, Ahmed SF, Alroughani R, Goadsby PJ. Migraine among medical students in Kuwait University. J Headache Pain. 2014 Dec 1;15(1):1–6.

24. Menon B, Kinnera N. Prevalence and characteristics of migraine in medical students and its impact on their daily activities. Ann Indian Acad Neurol [Internet]. 2013 Apr [cited 2020 Sep 21];16(2):221–5. Available from: https://pubmed.ncbi.nlm.nih.gov/23956569/

25. Perveen I, Parvin R, Saha M, Bari MS, Huda MN, Ghosh MK. Prevalence of Irritable Bowel Syndrome (IBS), Migraine and Co-Existing IBS-Migraine in Medical Students. J Clin DIAGNOSTIC Res. 2016;10(11):OC09.

26. Eysenbach G. Improving the quality of web surveys: The Checklist for Reporting Results of Internet E-Surveys (CHERRIES) [Internet]. Vol. 6, Journal of Medical Internet Research. Journal of Medical Internet Research; 2004 [cited 2020 Jul 19]. Available from: /pmc/articles/PMC1550605/?report=abstract

27. Mollayeva T, Thurairajah P, Burton K, Mollayeva S, Shapiro CM, Colantonio A. The Pittsburgh sleep quality index as a screening tool for sleep dysfunction in clinical and non-clinical samples: A systematic review and meta-analysis. Vol. 25, Sleep Medicine Reviews. W.B. Saunders Ltd; 2016. p. 52–73.

28. Lipton RB, Dodick D, Sadovsky R, Kolodner K, Endicott J, Hettiarachchi J, et al. A self-administered screener for migraine in primary care: The ID migraine™ validation study. Neurology [Internet]. 2003 Aug 12 [cited 2020 Jun 25];61(3):375–82. Available from: https://n.neurology.org/content/61/3/375

29. Cousins G, Hijazze S, Van De Laar FA, Fahey T. Diagnostic accuracy of the ID migraine: A systematic review and meta-analysis. Vol. 51, Headache. 2011. p. 1140–8.

30. Tfelt-Hansen P, Pascual J, Ramadan N, Dahlöf C, D’Amico D, Diener H-C, et al. Guidelines for controlled trials of drugs in migraine: third edition. A guide for investigators. Cephalalgia. 2012 Jan;32(1):6–38.

31. Stewart WF, Lipton R, Kolodner K, Liberman J, Sawyer J. Reliability of the migraine disability assessment score in a population-based sample of headache sufferers. Cephalalgia [Internet]. 1999 [cited 2021 Jun 25];19(2):107–14. Available from: https://pubmed.ncbi.nlm.nih.gov/10214536/

32. Oraby MI, Soliman RH, Mahmoud MA, Elfar E, Abd ElMonem NA. Migraine prevalence, clinical characteristics, and health care-seeking practice in a sample of medical students in Egypt. Egypt J Neurol Psychiatry Neurosurg [Internet]. 2021 Dec 1 [cited 2021 Jun 25];57(1):1–9. Available from: https://doi.org/10.1186/s41983-021-00282-8

33. de Almeida CMO, Lima PAM da S, Stabenow R, Mota RS de S, Boechat AL, Takatani M. Headache-related disability among medical students in Amazon: A cross-sectional study. Arq Neuropsiquiatr. 2015 Dec 1;73(12):1009–13.

34. Ferri-de-Barros JE, de Alencar MJ, Berchielli LF, Castelhano Junior LC. Headache among medical and psychology students. Arq Neuropsiquiatr. 2011;69(3):502–8.

35. Ojini F, Okubadejo N, Danesi M. Prevalence and clinical characteristics of headache in medical students of the University of Lagos, Nigeria. Cephalalgia. 2009 Apr;29(4):472–7.

36. Balaban H, Semiz M, Senturk IA, Kavakci O, Cinar Z, Dikici A, et al. Migraine prevalence, alexithymia, and post-traumatic stress disorder among medical students in turkey. J Headache Pain. 2012 Aug;13(6):459–67.

37. Alwahbi MK, Alamri MM, Alammar AM, Alanazi AM, Alotaibi AG, Alharbi SK, et al. Prevalence of Migraine among Medical Students of King Saud Bin Abdulaziz University for Health Sciences. Int J Sci Res. 2017;6(2):894–8.

38. Amayo EO, Jowi JO, Njeru EK. Headache associated disability in medical students at the Kenyatta National Hospital, Nairobi. East Afr Med J. 2002 Oct;79(10):519–23.

39. Seifert T, Sufrinko A, Cowan R, Scott Black W, Watson D, Edwards B, et al. Comprehensive Headache Experience in Collegiate Student-Athletes: An Initial Report From the NCAA Headache Task Force. Headache [Internet]. 2017 Jun 1 [cited 2020 Sep 21];57(6):877–86. Available from: https://pubmed.ncbi.nlm.nih.gov/28480575/

40. Birru EM, Abay Z, Abdelwuhab M, Basazn A, Sirak B, Teni FS. Management of headache and associated factors among undergraduate medicine and health science students of University of Gondar, North West Ethiopia. J Headache Pain. 2016 Dec 1;17(1).

41. Lebedeva ER, Kobzeva NR, Gilev D V., Olesen J. Factors Associated with Primary Headache According to Diagnosis, Sex, and Social Group. Headache. 2016 Feb;56(2):341–56.

42. Gupta S, Mehrotra S, Villalón CM, Perusquía M, Saxena PR, MaassenVanDenBrink A. Potential role of female sex hormones in the pathophysiology of migraine. Pharmacol Ther. 2007 Feb;113(2):321–40.

43. Gu X, Xie Y. Migraine attacks among medical students in Soochow University, Southeast China: a cross-sectional study. J Pain Res. 2018 Apr;11:771–81.

44. Panconesi A. Alcohol and migraine: Trigger factor, consumption, mechanisms. A review [Internet]. Vol. 9, Journal of Headache and Pain. BioMed Central; 2008 [cited 2020 Sep 22]. p. 19–27. Available from: https://thejournalofheadacheandpain.biomedcentral.com/articles/10.1007/s10194-008-0006-1

45. Rozen TD. A history of cigarette smoking is associated with the development of cranial autonomic symptoms with migraine headaches. Headache. 2011 Jan;51(1):85–91.

46. Aamodt AH, Stovner LJ, Hagen K, Bråthen G, Zwart J. Headache prevalence related to smoking and alcohol use. The Head-HUNT Study. Eur J Neurol [Internet]. 2006 Nov 1 [cited 2020 Sep 22];13(11):1233–8. Available from: http://doi.wiley.com/10.1111/j.1468-1331.2006.01492.x

47. Ibrahim NK, Alqarni AK, Bajaba RM, Aljuhani FM, Bally AM, Wakid MH. Migraine among Students from the Faculty of Applied Medical Sciences, King Abdulaziz University, Jeddah, Saudi Arabia. J Adv Med Med Res. 2018 Nov;27(11):1–10.

